# Moving from development to implementation of digital innovations within the NHS: myHealthE, a remote monitoring system for tracking patient outcomes in child and adolescent mental health services

**DOI:** 10.1101/2021.06.09.21257998

**Authors:** Anna C Morris, Zina Ibrahim, Omer S Moghraby, Argyris Stringaris, Ian M Grant, Lukasz Zalewski, Stuart McClellan, Garry Moriarty, Emily Simonoff, Richard JB Dobson, Johnny Downs

## Abstract

This paper reports our experience of developing, implementing, and evaluating myHealthE (MHE); a digital innovation for Child and Adolescents Mental Health Services (CAMHS) which automates the remote collection and reporting of Patient Reported Outcome Measures (PROMs) into National Health Services (NHS) electronic health care records. We describe the logistical and governance issues encountered in developing the MHE interface with patient identifiable information, and the steps taken to overcome these development barriers. We describe the applications architecture and hosting environment to enable it to be operable within the NHS, as well the as the capabilities needed within the technical team to bridge the gap between academic developers and NHS operational teams. We present evidence on the feasibility and acceptability for this system within clinical services and describe the process of iterative development, highlighting additional functions which were incorporated to increase system utility. This article provides a framework with which to plan, develop and implement automated PROM collection from remote devices back to NHS infrastructure. The challenges and solutions described in this paper will be pertinent to other digital health innovation researchers aspiring to deploy interoperable systems within NHS clinical systems.

## Background

Patient Reported Outcome Measures (PROMs) are recognised as indispensable clinical tools to document and enhance patient experiences, [1] and support outcome improvement. [1, 2, 3, 4] Implemented correctly, standardised outcome measures are an effective way to record self-reported changes in clinical outcomes, inform clinical practice and monitor service value. [1, 2, 3, 4, 5, 6, 7] However, long-term PROM collection is not part of routine practice in mental health clinics in the United Kingdom (UK). [8, 9] Particularly within Child and Adolescent Mental Health Services (CAMHS), survey findings show that guidelines for recording PROMs are adhered to in as few as 6-30% of clinical cases. [10, 11, 12] Furthermore, a study analysing the electronic health records (EHRs) of ADHD child and adolescent patients at the South London and Maudsley National Health Service Foundation Trust (SLaM NHS FT), one of the largest mental health institutes in Europe, found that less than 1% of longitudinal PROMs are recorded. [13]

Low rates of PROM recording, especially in CAMHS [14] have been attributed to patient and service level factors. [5, 11] Despite the known value of PROMS, demanding workloads, limited time, and resources mean that clinical teams are often too stretched to systematically administer and collect these questionnaires and tend to rely on impressions to record patients’ progress. [15] Low response rates from families to structured outcome measures and lack of feedback available to both clinicians and patients have been found to be contributing factors. [16] In response, online electronic PROM (ePROM) systems have been designed and provisioned to support outcome monitoring in healthcare settings by offering a practical solution to identified barriers of collecting paper-based PROMs. [17, 18] Moreover, enthusiasm for web-based monitoring portals has increased rapidly in the current climate due to the emergent COVID-19 pandemic, rendering health care providers under mounting pressure to offer services online. [19]

Despite the growing need and support for ePROMs, most existing platforms target the clinical management of physical health, [20, 21, 22, 23, 24] rather than mental health, where the success of these tools is varied. [25, 26, 27]. Furthermore, the benefits of remote monitoring platforms may be limited by their functionality. For example, while a recent review of ePROM collection tools identified 33 exemplars in the field of clinical oncology, only 37% of these allowed respondents to complete PROMs outside the clinical setting i.e., from their own home, and just 44% of remote monitoring tools had the capacity to integrate collected data directly to patient’s EHRs. [28] Moreover, electronic platforms often rely on input from clinicians to set the type and frequency of follow-up outcome measures required, [28] thus limiting the potential time and effort savings projected from ePROM delivery. [29] At present, the majority of ePROM system are developed as standalone platforms, independent of the information technology (IT) infrastructure responsible for handling routinely collected health information, therefore, patient consent is normally required before ePROM systems can be used to request clinical information at the patient-level. [30]

Finally, notwithstanding the promise of technology to transform the way the National Health Services (NHS) collects stores and displays clinical information, digitalisation is purported to develop at a slower rate relative to other health organisations or industries. [31, 32, 33] This suggests that cultural factors within the NHS, as well as the mechanics of developing e-platforms, may disproportionately affect the organisation’s efforts to improve healthcare by introducing paperless strategies.

Our experience commenced with the development of APPROaCh (Agent Platform for automating patient PROvided Clinical outcome feedback), [34] a distributed multi-agent framework enabling the provision of remote, proactive, and personalised collection and assessment of PROMs without clinicians’ intervention.

In this paper, we aim to: (1) provide a reflective account on our experience in the development, deployment, testing and evaluation of myHealthE (MHE), adapting the APPROaCh platform to target CAMHS in the NHS of the UK, specifically within SLaM; (2) describe the resulting system in relation to its technical architecture, data flows, and human input, and; (3) highlight the key barriers of development and how these could apply to other novel health innovations seeking to deploy within NHS infrastructure.

We provide a description of the interdisplinary approach we took to build a cloud-based environment that can host an NHS digital health monitoring system (DHMS) to permit the safe processing and storage of clinical data. We describe the key information governance and IT clearance processes required to ensure adequate data protection, resulting amendments to existing system architecture and barriers posed by security issues relating to service user involvement. To our knowledge, this is the first online management system described in the literature, that is compatible with NHS EHRs and automates correspondence, delivery, and collection of electronic PROMs at predefined post-treatment intervals to address the low return rates of primary carer outcome measures in UK CAMHS, in accordance with National PROM collection guidelines. [35] We describe the processes undertaken to acquire adequate clarity around ownership and policy relating to maintenance and responsibility. We outline steps taken to bridge the gap between frontline care and innovation research and raise questions around how to support future digital initiatives in the NHS. Finally, we provide an implementation strategy which helped us ensure that clinical service support was developed and maintained throughout.

## Methods

### myHealthE Team

The interdisciplinary system development team was led by a King’s Health Partnership group combining academic and clinical expertise from King’s College London (KCL) and SLaM NHS FT. A university-based research team, comprising a clinical academic lead (JD) and research assistant (AM), was responsible for daily project management and implementation, and worked closely with KCL-employed health informaticians (ZI, IG, LZ) and SLaM Information governance, digital clinical systems leads (SM, GM) to prepare MHE for clinical use.

### Generic myHealthE technical architecture

MHE achieves repeat, automated follow-up patient progress tracking via the integration of two primary components: (1) a multi-agent tracking program, which is comprised of a subset of agents adapted from those previously described - [34] primarily including the agents DataManager, UnitManager and PatientFollowUp - and an associated database. This software monitors internal patient health records, imports patients’ existing baseline questionnaire responses, enrols them in the system, and creates a patient-specific schedule for notifying the caregiver to complete follow-up measures; (2) a patient-facing web application which allows the caregiver to log in and fill out online questionnaire forms to log new measures, to a schedule that is pre-determined by the tracking agent. These additional questionnaire results are logged into the same database that the tracking agent interfaces with, thus creating a feedback loop aligned to the patient-specific schedule originally determined by the tracking agent.

### myHealthE Development

#### Establishing system specifications

Initial specification requirements were developed by the MHE team and focused on three main themes: user experience, security, and scalability. Early examples of MHE specification requirements included the need to identify a (cloud-based) server suitable for hosting the application and patient identifiable information (PII), a reliable and secure text and email communication mechanism, and a safe method of caregiver registration verification. Having developed MHE from an NHS governance and engineering perspective, we contracted a local commercial enterprise specialising in website and application development to help design an engaging and user-friendly patient interface.

#### Proposed system data flow and technical components build

Figure 1. illustrates the MHE extraction and imputation data flow process. To align with hospital standards for collecting and managing patient identifiable information, we deployed the system on a virtual server that was provisioned within SLaM’s Microsoft Azure cloud subscription; this provides direct, private integration with SLaM’s IT infrastructure (including the EHR), protected by the same institutional firewalls and relevant cybersecurity protocols as the rest of the SLaM digital estate.

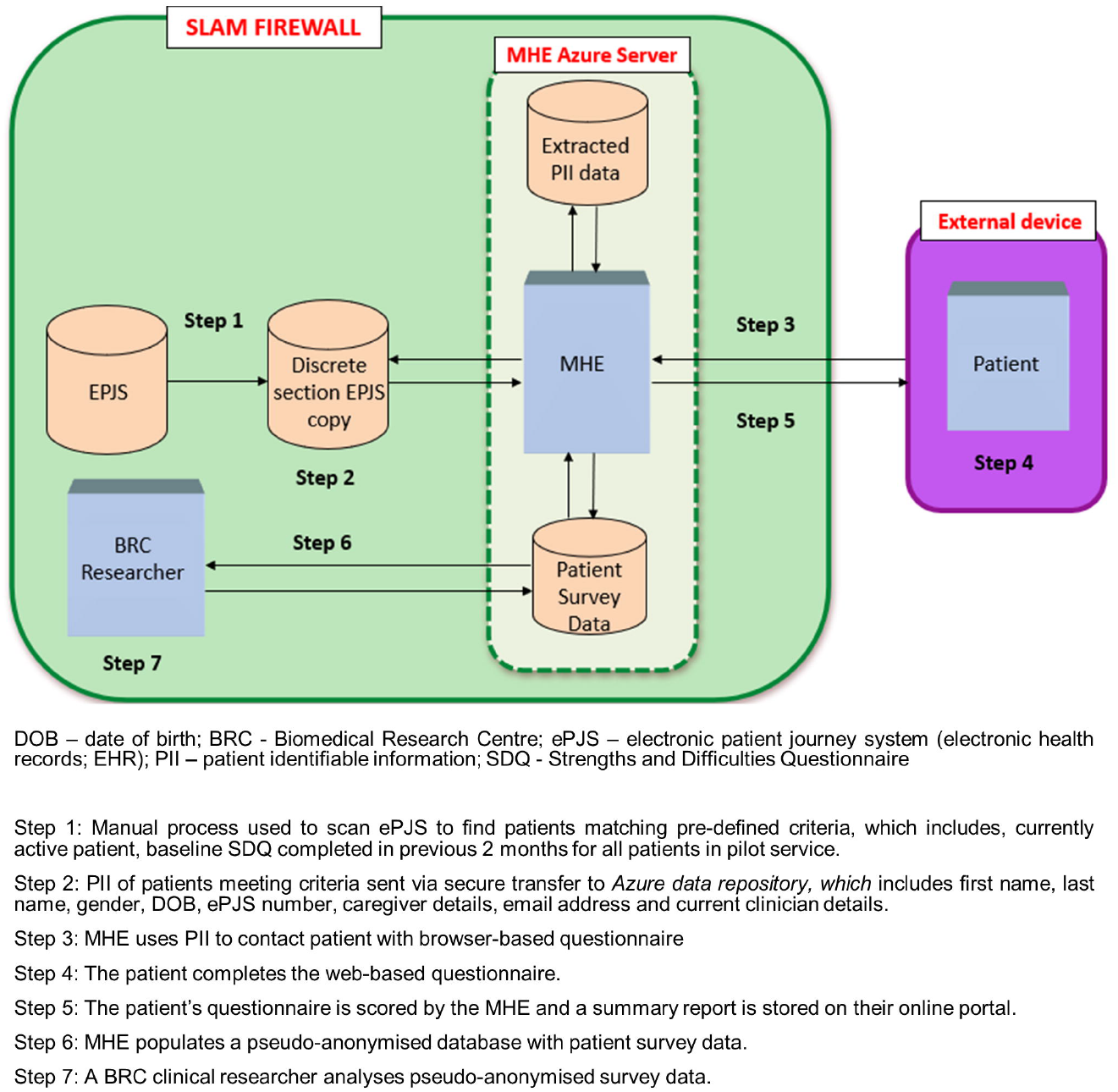

The prescribed SLaM server hosts two distinct databases which are populated via the MHE agent framework. Firstly, a SQL patient identifier database is populated through an MHE-specific script (implemented by the SLaM Systems Team) in SLaM’s EHR (also known as the electronic Patient Journey System; ePJS). The script extracts child and caregiver patient identifiable information (PII); these include first name, last name, sex, date of birth, and contact details (e.g., mobile numbers and email addresses) for specific patient populations. Selected children include active CAMHS patients who require a follow-up, parent-reported Strengths and Difficulties Questionnaire (SDQ-P) [36] to be completed (patients who have a completed baseline SDQ-P with no follow-up SDQ-P entered). Secondly, a pseudo-anonymised dataset populated with patient survey data, generated through patient caregivers completing online SDQ-P measures. MHE also provides a mechanism for the Trust to know which caregivers are happy to be contacted about the prospect of enrolling in future Trust-led research studies – a consent for research contact register which will be stored in this database. Trust approved researchers have access to patient reported outcomes

### Anticipated translational challenges

After determining prerequisite application architecture and data flows, the interdisciplinary group identified several potential barriers which could restrict the development and transfer of MHE from a research environment into clinical practice.

#### 1. Legacy Systems

Electronic Healthcare Record and Digital Systems (especially in the UK) have largely comprised of specialised proprietary technologies. As with many NHS Healthcare providers, SLaM EHR software systems were not designed to be interoperable, nor remotely accessible, but built for meeting the immediate clinical and clerical needs of recording and storing captured data from patient interactions. Building innovations that can enhance the capabilities of these proprietary systems within the NHS, has been hampered by the lack of centrally agreed data standards for sharing, or open software systems for shared development. With no mandate for interoperability, it has been historically difficult for NHS organisation to encourage EHR and technology suppliers to adopt DHMS from research into clinical practice. [37]

#### 2. Introduction of cloud-based solutions

Recent guidelines published by NHS Digital (2018) recommend that all NHS services transition from using locally managed servers to store patient information to public cloud-based solutions. This recommendation releases NHS organisations from some of their maintenance obligations to continually improve and upgrade local systems and permits the development of digital products that are not constrained by data storage or processing limits. Because of the recent change in guidance, most NHS organisations are yet to establish their own in-house standards and processes for developing DHMS cloud-based environments suitable for safe processing and storage of clinical data.

#### 3. Data security and information governance

MHE was developed to replace a pen and paper method for collecting sensitive and personal clinical data. The security parameters for our application needed to provide full system protection, in line with SLaM’s policy for existing electronically held patient records. New relationships would need to be established between KCL and Trust’s governance and IT personnel, to enable clear communication throughout the project phases to enable testing, evaluation, and deployment of MHE as a digital innovation.

#### 4. Resource barriers

At an organisational and system-wide level the NHS was not financially structured to support innovation design, delivery, or dissemination in addition to existing workload. [38] We expected to encounter project delays during the development stage owing to a lack of designated organisational personnel to oversee project management and construction of innovative digital technologies. [39, 40]

#### 4. Product ownership

MHE was created in a university research and development (R & D) environment as a system to enhance routine clinical practice. The business case had yet to be fully established for SLaM to adopt this innovation to be maintained as business as usual, as with all other clinical systems. Hence MHE would require support and resources from already stretched SLaM services, as an addition to their existing workloads.

#### 5. Lack of engagement from clinical team

Enthusiasm within the clinical service selected for MHE implementation could diminish if protracted technical delays were encountered over the course of deployment.

### Implementation setting

The study was conducted at Kaleidoscope, a community paediatric and children’s and young people’s mental health centre based in Lewisham, South London within the Lewisham Neuro-developmental team (NDT), between the 11th of February 2019 and the 14th of May 2019.

### Implementation strategy

Too often, innovations in digital health fail to reach frontline care owing to poorly defined implementation strategies. Adoption of electronic systems into established clinical settings is a complex process, reliant on several important multi-level innovation and organisational factors. [41] Applying lessons from health informatics implementation literature and the Consolidation Framework for Implementation Research (CFIR) framework, [3, 25, 42] MHE introduction followed a staged approach, designed to alleviate any security concerns, and foster local ownership of the system. This approach consists of four pre-implementation stages and is anticipated to take six months to complete.

#### 1. Baseline evaluation

To provide a baseline assessment of caregiver reported SDQ completion rates for SLaM CAMHS, we will run searches through Clinical Records Interactive Search (CRIS), a database comprised of de-identified records for all patients accessing Trust services. [43, 44, 45] This data will allow us to assess whether our automated electronic system outperforms current SLaM SDQ-P data collection procedures.

#### 2. Orientation

This stage will focus on site preparation, achieved through regular team consultations at clinical team meetings. Planned sessions will be used to discuss the aims and expectations of the project, assess local capacity for MHE adoption and develop fundamental communication channels within the service.

#### 3. Stakeholder engagement

Stakeholders will be familiarised with MHE functionality and given the opportunity to provide system feedback through separate staff and parent group interactive demonstrations of a prototype MHE system.

#### 4. System refinement

Understanding that the application meets stakeholders’ needs is imperative to the success of the application, therefore, feedback from the stakeholder engagement stage will be used to refine the MHE prototype ready for application and to establish joint expectations for implementation protocol.

## Results

### Data security and information governance

To identify and prevent privacy breaches, new ICT initiatives developed in the NHS environment for use with PII are required to undergo a data privacy impact assessment (DPIA). Following an initial security review, the web-application server hosting the MHE prototype failed to meet privacy protection standards. A replacement web-application server within the Microsoft Azure (MS Azure) cloud platform was identified as an appropriate host for the multi-agent system. Following subsequent MS Azure DPIA approval, funding was secured to provision two MHE specific internal servers and ensure appropriate data protection and system security features were built. Penetration testing on the agent code and data flow connectivity were outsourced to a SLaM endorsed industry partner. All password protected data repositories are located within the SLaM firewall and a disaster recovery strategy has been developed for system restoration in the event a critical issue is encountered.

### myHealthE application profile refinements

Overall, the new MHE environment build was sustained for 23 months. The internal Azure setup is provided in Figure 2.

#### Critical pieces

A web application frontend (patient/user-facing web site), web application backend (CMS and database interface for the frontend) and tracker agent (Java-based ePJS interface, scheduler & notification dispatcher);
Core system components - web server (Apache, inc. PHP, serves web frontend & backend), application database server (local MySQL instance, hosts backend CMS & logins, and SDQ/tracker data) and Azure server instance (CentOS Linux server, runs above components);
External dependent services - Gov.UK notification service (service/tracker emails and SMS), Cloudflare network (traffic routing, TLS certificate), SendGrid mail service (password reset emails) and ePJS database (if new patients/SDQs need to be ingested).

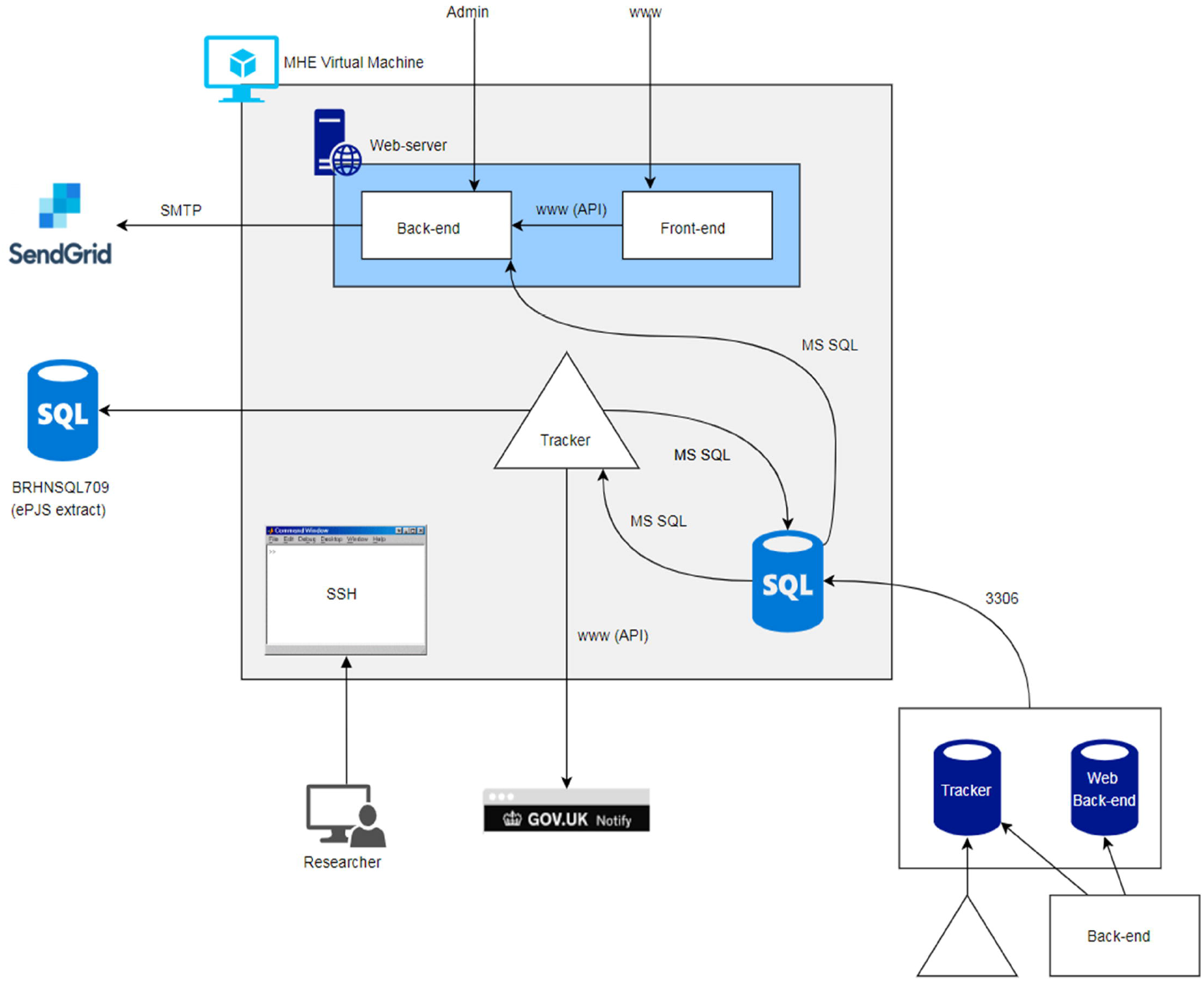

To make information collected via MHE available for potential clinical use, we aspired to enter this data directly into SLaMs main electronic patient records system – ePJS. However, initial scoping revealed that ePJS does not provide an API to allow this data transfer to occur. During development the MHE team were approached by the Trust’s clinical systems team with an opportunity to trial a novel data processing tool – Robotics Processing Application (RPA), which would enable this. In brief, RPA runs visual basic scripts (VBS) programmed to simulate data entry through the front-end of patient’s electronic health records i.e., logging into ePJS, entering patients’ unique identifiable Trust ID, selecting active episode of care and inputting data points. A six-week proof of concept is currently underway to trial the process. To date, the RPA process has been trialled on both the test and live ePJS platforms, resulting in successful data transfer for new SDQ data collected via MHE.

### Feasibility testing

Informed by pre-development exploration with caregivers about the acceptability and practicability of a DHMS for supporting their child’s treatment; the MHE team worked iteratively with front-end application developer to re-design the MHE website. After initial development, necessary adjustments and design changes were made to the platform on a retainer contract basis meaning platform appearance was adaptive to issues identified through comprehensive within-team testing.

The resulting prototype was thoroughly tested by the MHE team throughout development using dummy login credentials. Testing was first conducted on the high performance and research cloud computing platform, *Rosalind*, which is hosted by KCL, and is co-funded by and delivered in partnership with KCL and the National Institute for Health Research (NIHR). It was subsequently moved to a live MHE staging platform located behind the SLaM firewall, to enable full ecological testing, which included inputting of new SDQ data, account registration verification, and website access via an extensive range of operating systems and internet-enabled devices.

### Development barriers

At the time of development, no firm guidelines were in place for implementing new digital innovations into SLaM NHS infrastructure. Clarity regarding SLaM’s product development procedures was obtained step-by-step with assistance from SLaM advisory groups, individuals on Centre for Translation Informatics (CTI) operations board (a research partnership jointly led by SLaM and KCL aspiring to improve healthcare using digital innovations), Project Management Office, and SLaM Information Governance personnel. Accordingly, we spent a substantial amount of time becoming versed in the time, governance, logistical and technical resources, and funds required to implement and sustain projects within SLaM IT infrastructure as described in Figure 3. We worked closely with SLaM IT’s operations department to achieve the resulting MHE technical build. However, due to the sensitive nature of the data being collected and processed by MHE it was difficult to deliver on our time sensitive grant funded milestones owing to SLaMs understandable need to manage business as usual and innovative projects simultaneously within their existing workforce. Therefore, despite having the in-house KCL technical skills and resources, identify, monitor, and resolve inevitable bugs, we were reliant on SLaM dedicated technicians to make changes to firewalls, Content Delivery Networks and ports to support its development and maintenance.

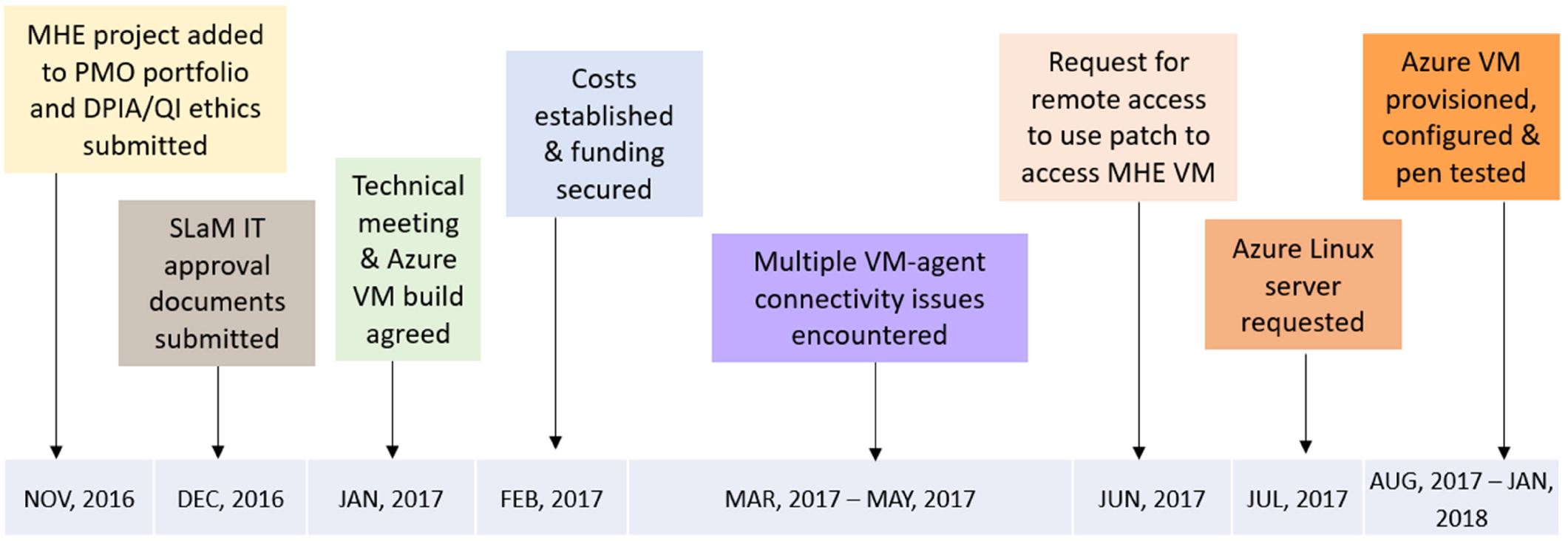

### myHealthE implementation

#### Baseline evaluation findings

We used CRIS to establish current levels of parent-reported SDQ collection for the participating service as well as all other child mental health services served by SLaM. Of the 28,382 CAMHS service user’s records surveyed, baseline SDQ-P were observed for approximately 40% (n=11,212) of the sample and of these cases only 8% (n=928) reported follow-up SDQ-P in subsequent 6 months. [14]

#### Orientation, stakeholder engagement and system refinement

Site participation was agreed through professional links between the research team clinical lead and service managers. AM attended routine clinical team meetings fortnightly, where project aims, and expectations were established. These visits were also used to provide real-time updates on product development. Delays in server development and application-server connectivity meant that this stage continued beyond the allocated six-month period. Professional links were maintained through email updates and clinic visits from AM, though visit frequency was reduced during this extension to enable more meaningful progress presentations.

Accordingly, stakeholder engagement activities commenced before a complete MHE prototype was available. Using an earlier prototype of MHE planned sessions were used to collect stakeholder feedback. In sessions with clinical staff AM provided a dummy run-through of the MHE portal from a user-perspective which provided staff with an idea of frequency and content of MHE communications caregiver’s, web-portal aesthetics, SDQ data entry requirements and response output. Through transparent open-ended discussion, staff raised initial questions and comments about procedural differences between automated data collection and the current distribution and collection of baseline and follow up SDQ-P forms and an overall positive regard for the proposed introduction of this system into routine care. Concerns centred on remote technology detracting from the therapeutic process through 1) limiting opportunity to use in session SDQ-P administration as a springboard to identify underlying concerns that may not be disclosed 2) caregivers’ potential to misunderstand questions or misinterpret SDQ-P output which could be negatively fed back to the patient and; 3) creating a lack of caregiver motivation due to online completion being perceived as a tick box exercise.

We were unable replicate the clinical staff user-testing sessions with caregivers, due to time issues. However, the same process was applied to a convenience sample of caregivers (n=3) on an individual basis who agreed to meet with AM following a clinical appointment to assess caregiver perspectives on acceptability and practicability of MHE for support their child’s treatment. Feedback was very positive and mainly centred on the clear layout of the website, trusted NHS branding and the anticipated ease of completing routine outcome measures. Once a working prototype of the current MHE platform was available, security issues associated with the MHE NHS firewall protected server configuration delays continued to impair our ability to invite patients and caregivers to experiment with the platform outside of the clinic to obtain ecologically valid feedback.

#### Trial initiation and preliminary findings

Following final system changes, all caregivers of active Lewisham NDT patients were contacted by letter to inform them of potential changes to the way Lewisham CAMHS gather clinical information about their patients (i.e., electronic rather than paper questionnaires) and contact information should they have any questions about the initiative. We witnessed a high turnover of staff within the Lewisham NDT service during the protracted orientation stage, rendering new team members less familiar with the trial and its purpose. Therefore, we decided to perform the trial using a single-blind study design meaning that clinicians would be unaware as whether their patients were receiving MHE monitoring or standard care to minimise any undue influence of clinician’s behaviour, i.e., encouraging MHE use on user engagement. The Lewisham clinical lead was contacted two weeks prior to the trial start date to obtain final approvals. The trial ran for twelve weeks between 11th of February 2019 and the 14th of May 2019. A total of 196 families, with at least one previous SDQ-P completed were enrolled to the trial (MHE provided n = 98 and care as usual provided n = 98). Seventy percent of caregivers who received the MHE filled out a minimum of one SDQ-P during this period compared to 8% of caregivers who continued with standard CAMHS care. Over the three months 87 follow-up SDQ-Ps were recorded via the platform. All accept one caregiver who registered on their personal MHE account continued to complete an electronic SDQ-P. Integration of MHE was mainly positive with the exception of one identified barrier, where caregiver contact details were only present in the expected area of ePJS for over half the patients enrolled to received MHE. This meant that the AM had to screen other structured or free text fields to identify contact information and enter it directly to the MHE back-end.

## Discussion

This paper describes implementation of state-of-the-art concepts in informatics research bearing immediate translational benefits to clinical care in mental health. We document the rational and methodology-supporting development of the MHE system within modern NHS technical infrastructure and key barriers surmounted to successfully deploy MHE within clinical systems.

### Added value of MHE

PROMs offer enormous potential to improve the quality of mental health service. [46] In MHE, we have designed and provisioned an online monitoring tool fully integrated with NHS electronic records to automate the collection of patients reported clinical information at pre-defined repeated post-treatment time points and to facilitate associated patient communication electronically. While electronic systems have demonstrated considerable promise to facilitate PROM uptake, [47, 48, 49] current examples of clinical value are more readily reported for physical health. [21, 48] Moreover, while successful implementation of eHealth platforms into existing health records has been reported in adult health [26] to our knowledge MHE is the first of its kind to achieve this in UK child mental health services.

In our feasibility trial, we observed levels of caregiver SDQ completion, which are substantially higher than rates reported for current paper-based practices. These findings suggest that MHE has the potential to tackle the time limiting step of documenting PROMs in CAMHS by allowing technology-supported remote outcome measure follow-ups and enabling effortless availability of reported information to patients treating care team. As such, MHE may improve transparency of care, by automating patient involvement in the process of collecting audit data and the efficiency and quality of care, by ensuring that clinicians are aware of their patients’ progress. Moreover, given the rapid shift to remote service delivery brought about by the COVID-19 pandemic and that the NHS has published an ambitious plan to digitalise the health sector over the next 10-years, [50] it is vital that paperless health monitoring innovations outperform current practice in a way that is safe and agreeable to patients and their health care providers.

This paper has highlighted how hard implementation efforts are. Even with the best laid out plans, delays in other elements of the project combined with the need to complete the implementation cycle within a 24-month charity funded budget time window had a knock-on effect for site preparation and majorly influenced our ability to involve families in the design of the patient portal. This is problematic since patient portal engagement is strongly influenced by the patient’s interest and capacity to use web-based portals. [51] Early indications from our feasibility trial suggest that the current platform was acceptable to families; end-user follow-up will be conducted to assess the acceptability of this current platform and how it could be improved to increase MHE engagement rates.

### Sustainability and scalability challenges

Following the deployment of MHE in a clinical environment, interest has been expressed in scaling up the system for use across SLaM CAMHS. Capacity issues within SLaM IT made it uncertain how long response times would be when faced with inevitable bugs and technical difficulties. CTI dedicated personnel were hired in an attempt to ease the burden of MHE development (and other CTI projects) placed on the digital services team who share R & D and operational responsibilities, which notably helped communication between product developers and SLaM IT. However, continued collaboration between research and IT teams is needed to establish how this role can further streamline the development-test-production pipeline. For example, providing trained CTI staff with the authority and operational capability to action approved changes from SLaM’s IT operations change acceptance process. Adjustments to firewalls, network and port changes are examples of operations that can hinder development progress, which could have a considerable impact on service user engagement, and likely dampen clinical service enthusiasm for being an implementation trial site.

A key technical challenge will be maintaining MHE constituent component harmonisation once rolled out across the Trust, to ensure uninterrupted data flow functionality. Monitoring insights were set up by the research team and CTI throughout development, however, the responsibility for checking these reports for each of the systems critical components and performing system maintenance i.e., software updates, will fall under the remit of SLaM IT once the system is added to the hospitals business-as-usual monitoring portfolio. Therefore, careful planning is needed to document and agree these procedures ahead of a full-scale launch to ensure adequate resources are available to support system amendments, advancements and restoration in a timely manner.

### Planned system expansion and application

Valuable findings from our feasibility trial highlighted ways the system could be improved, for example, the difficulties experienced extracting caregiver contact details from structured clinical records, which could be remedied by manualising caregiver contact information entry upon MHE registration or allowing MHE to search multiple EHRs locations by refining the SQL scripts responsible for this process or using natural language processing (NLP) methods to identify this information in free text records. Plans are in motion to expand the MHE framework to increase functionality, including referral tracking, information gathering and signposting features as well as the addition of condition specific outcome measures and adult PROM questionnaires to facilitate mental health service delivery more broadly. Funding has also been secured to interface with other technologies capturing neuropsychological and accelerometer data and online intervention delivery platforms. Now we are equipped with the knowledge and technical support to build and deploy solutions in NHS-walled servers’ phase two development should be less time consuming and allow more time for planned theoretically informed end-user testing.

### Conclusion

We provide a worked example of multi-agent platform development to improve patient reported outcome collection using timely personalised communications to guide self-care outside the hospital environment, strengthen families sense of support and increase their commitment to treatment. Such frameworks provide a cornerstone in the applicability of digital health outcome monitoring research, a relatively young field with large potential but few real-world applications in mental health clinical practice. By overcoming barriers to operate within NHS clinical systems MHE can automate routine clinical data collection which are infrequently reported in CAMHS. The system will ease clinician burden and provide a bridging connection with their patients using methods beyond the scope of current clinical practice.

## Data Availability

Data are available on reasonable request. The data accessed by CRIS remain within an NHS firewall and governance is provided by a patient-led oversight committee. Access to data is restricted to honorary or substantive employees of the South London and Maudsley NHS Foundation Trust and governed by a local oversight committee who review and approve applications to extract and analyse data for research. Subject to these conditions, data access is encouraged and those interested should contact, CRIS academic lead.

## Declarations

### Ethical approval

This study was approved by the SLaM NHS Foundation Trust Quality Improvement and Service Evaluation Ethics panel.

### Guarantor

JD

### Contributorship

ACM (corresponding author), ZI and JD researched literature and coconcieved this study. ACM and ZI co-wrote a first draft of the manuscript. ACM and JD revised and finalised the manuscript. OSM, IMG, LZ and SM supported study design. GM supported study design and data acquisition. All authors reviewed and provided critical revisions to the manuscript and approved the final version of the manuscript.

## Acknowledgements

The authors give thanks to the MHE digital development team Digital Marmalade, with particular thanks Andy McEniry and Jeremy Jones. The authors acknowledge use of Clinical Records Interactive Search (CRIS), which is supported by the NIHR Biomedical Research Centre for Mental Health (BRC) Nucleus at the SLaM NHS Foundation Trust and Institute of Psychiatry, Psychology and Neuroscience (IoPPN), King’s College London (KCL) jointly funded by the Guy’s and St Thomas’ (GSST) Trustees and the South London and Maudsley Trustees. The authors acknowledge use of the research computing facility at KCL, *Rosalind* (https://rosalind.kcl.ac.uk), which is delivered in partnership with the NIHR Biomedical Research Centres at SLaM and Guy’s & St. Thomas’ NHS Foundation Trusts, and part-funded by capital equipment grants from the Maudsley Charity (award 980) and Guy’s & St. Thomas’ Charity (TR130505). The views expressed are those of the author(s) and not necessarily those of the NHS, the NIHR, KCL, or the Department of Health and Social Care.

